# Perspectives on using a celebrity narrative to teach the psychiatric formulation to final year medical students

**DOI:** 10.1101/2023.12.07.23299651

**Authors:** David Hickey, John McFarland

## Abstract

**Objectives:** The proficiency of an accomplished psychiatrist encompasses the development, adaptation, and refinement of a biopsychosocial formulation. A formulation aids clinical decision-making by organizing information and furnishes a documented rationale for decisions. Despite its significance, psychiatric trainees often perceive the formulation as arduous and time-intensive, leading to avoidance. Educational shortcomings are pervasive. Addressing this calls for low-cost, novel solutions. Undergraduate medical education possesses aspiring psychiatrists and provides a platform to develop a foundational understanding of formulation. This study aims to explore the viability of a novel, resource-efficient approach employing a celebrity case narrative to attain a fundamental understanding of the psychiatric formulation while concurrently elucidating activated pedagogical cognitions by using such methodology from the student’s perspective.

**Methods:** A psychiatric formulation tutorial was conducted across five distinct final-year medical student cohorts during one academic year. A standardized tutorial structure was devised, incorporating interchangeable case studies. Following the tutorial, a post-tutorial survey was administered, followed by interviews that underwent qualitative analysis.

**Results:** Seventy-seven participants responded to the survey, expressing favourable views. Twenty students consented to interviews. They were distributed across five sessions with an average of four participants per group. Interviews yielded five key themes: Understanding of formulation, cognitive engagement, emotional salience and ethical considerations with twelve corresponding subthemes. The results suggested viability in using this methodology to teach formulation to foster a basic understanding and elicited a range of pedagogical phenomenon that may have contributed to this understanding.

**Conclusion:** This study indicates that integrating a celebrity narrative into psychiatric formulation teaching intervention bears potential for enhancing engagement concurrently mitigating negative attitudes towards the formulation. The approach reveals latent learning outcomes suggesting a profound pedagogical impact. The range of pedagogical process elucidated lays a foundational research base for future instructional design and research.

**Declaration of interest:** None

## Introduction

### The psychiatric formulation

#### Background

This study focuses on the biopsychosocial formulation, specifically the ‘4 P model’ (Bolton, 2014). Originally conceptualized by Engel (1977) and refined from Meyer’s multiaxial foundations (1951), it is deemed crucial in psychiatry and “*basic to daily clinical practice”* (Sperry, 1992). As a multidimensional diagnostic tool, it encompasses biological, psychological, social, and behavioural aspects. The formulation is an exercise in clinical reasoning that helps the writer to think clearly about the diagnosis, aetiology, treatment, and prognosis (Harrison, 2018). It provides a person-centred cross-sectional view of the patient’s present condition, considering historical influences and ultimately exploring why the patient is “*unwell in this way, at this time*” (Meagher et al., 2020). In contrast to biomedical models like the International Classification of Diseases (World Health Organization, 2019), the formulation is multifactorial and holistic. It complements rather than competes with traditional biomedical diagnostic models.

#### Educational shortcomings

Literature pertaining to psychiatric formulation teaching for undergraduate medical education is sparse. Postgraduate psychiatry trainees represent the closest comparable cohort with an established literature base. Among trainees, widespread deficiencies in formulation education are prevalent and probable barriers have been considered (McClain et al., 2004: Ratanawongsa et al., 2006). Trainee cohorts have recognised their own deficiencies and underscored the necessity for enhanced formal instruction in formulation (Crockford et al., 2004). The near total lack of compliance among trainees in documenting formulations within admission reports in a UK institution suggest shortcomings may have lasting clinical implications (Abbas et al., 2013).

Perceived difficulty has been identified as a potential major contributor to shortcomings; in a resident cohort from the USA, formulation was rated as the second most challenging skill, only behind psychotherapy (Sullivan et al., 2020). An Australian review posited that imprecision, subjectivity, and multifactorial vertical integration render formulation an intimidating task for residents (Bagster et al., 2021). Logically, the linear consequence of negative attitudes is evasion, inadequate skill attainment, insufficient skill mastery and subsequently a depletion of skilled educators perpetuating shortcomings. The absence of standardization across training institutions has confounded deficiencies (Ben-Aron & McCormick, 1980: Fleming & Patterson, 1993: McClain et al., 2004). Similar barriers likely exist in medical student cohorts in conjunction with competing educational opportunity costs.

#### Educational interventions

Various educational strategies encompassing resource allocation have undergone investigation. Time allocation for interventions varies in these studies, ranging from one-session seminars (Guerrero et al., 2003), one-day seminars (Sullivan et al., 2020), to 12-week programs (Ross et al., 2016). Methodology of intervention varies greatly from minimal instructional design such as raising awareness of clinical need and existing shortcomings (McLain et al., 2004), to precise and targeted curricular planning (Ross et al., 2016). Furthermore, innovative approaches diverging from conventional didactic teaching models have been explored including utilising visual metaphors (Alyami et al., 2015), pattern-based learning (Fernando et al., 2012) or character exploration within feature films (Mish, 2000).

The predominantly favourable outcomes of these studies indicate that the complexity of skill acquisition might not inherently underlie educational deficiencies. Instead, attitudes and cultural factors could influence perceived difficulty, potentially resulting in task avoidance and degradation of skills over time (Bagster et al, 2021) (Smith, 2021). Consequently, it is reasonable to suggest fear and a fundamental lack of foundational comprehension regarding clinical benefits are significant barriers to education around formulation. An opportunity thus presents itself to explore the potential of a novel approach to learning to foster core understanding early in an aspiring psychiatrist’s career and to cultivate positive attitudes.

### Celebrity culture

#### History of celebrity

The innate fascination with celebrities transcends the confines of contemporary society. Indications point to the emergence of modern celebrity during the 18th century, a period marked by the capacity to widely disseminate names and visages through the advent of the printing press (Gamson, 2004). An extensive body of literature and artefacts originating from ancient Greek and Roman civilizations suggests that the notion of celebrity, or a concept akin to it, existed in some form well before the 18th century (Garland, 2010). The abundance of modern and historical figures with universally recognised narratives offers a substantial wellspring of communal knowledge and intrigue.

#### Psychological intrigue of celebrity

The underlying motivations fuelling human attraction to celebrities remain enigmatic. Barkow (1992) postulated that the proliferation of parasocial relationships facilitated by mass media has activated an evolutionarily ingrained psychological impetus for acquiring social information. It is plausible that celebrities, both unintentionally and perhaps deliberately, tap into humanity’s innate drive for social interaction, thereby infiltrating an individual’s distinctive social network, social circle, or even perceptions of kinship (Dunbar & Spoors, 1996). The private sectors have capitalised and profited on these psychological processes for some time (Redmond & Holmes, 2007; Zafer, 1999).

#### Academic uses of celebrity narratives in mental health

The utilization of celebrity narratives as an academic pursuit within the field of psychiatry is not novel. Goldberg (2020) constructed an opinion piece that outlines suitable case studies to teach about clinical formulation. Kalb (2018) authored a book exploring mental illness in a range of diverse public figures. A plethora of further publications delves into historical and contemporary ‘celebrity’ figures indicating that this academic intrigue transcend temporal and thematic boundaries. Consider, for instance:

- Monarchs: The linguistic analysis of King George’s purported mania, along with an investigation into King Charles VIII’s alleged brain injury (Rentoumi et al., 2017; Zaenllo et al., 2021).
- Classical Musicians: Multitudes of publications explore the potential mental disorders afflicting Mozart (Karhausen, 2010).
- Post-Impressionist 18th Century Painters: A retrospective psychoanalysis of Vincent Van Gogh utilizing diverse sources and expertise (Nolen et al., 2020).
- 20th/21st Century Actors: An examination of Jack Nicholson’s contributions to neuroscience and a discussion on Robin Williams’ suicide (Tripathi et al., 2017; Tohid, 2016).
- Novelists: A psychological autopsy of Ernest Hemingway (Martin, 2006).

The utilization of celebrity profiles as an educational tool transcends the boundaries of the psychiatric specialty. It is evident that an inherent human captivation with celebrity profiles exists, and this has been harnessed for scholarly pursuits within psychiatry for decades. However, what remains lacking is a foundational understanding of the processes underpinning the use of these figures for educational endeavours. A qualitative analysis of learners’ perspectives is of significant value to establish a knowledge base that enhances comprehension of the learning mechanisms inherent in leveraging this potentially valuable educational resource.

### Research focus

The principal aim of the current study is to investigate the viability of using a celebrity narrative case study to cultivate a core understanding of the psychiatric formulation from final year medical students’ perspective. In conjunction with this, the qualitative design allows for further exploration into the advantages and disadvantages of using this methodology and allows for theoretical consideration of the provocation or evolution of pedagogical learning processes.

## Methods

### Context

The study participants were in the final year of their postgraduate medical school program at the University of Limerick’s School of Medicine during the academic year 2022/2023. All students were encouraged to attend the formulation tutorial, which was integrated into the standard curriculum for the first week of their psychiatry rotations. Participation in the study was voluntary. Attendance of tutorial and participation in the study was not mutually exclusive.

### Ethical approval

Ethical approval for the involvement of medical students was obtained from the University of Limerick’s Research Ethics & Governance. The project ID is 2022_12_15_EHS.

### Structure of the tutorial

A psychiatric formulation tutorial was conducted for five distinct participant groups throughout one academic year. The tutorial structure was standardized (see Table 1), with the only variation among participant groups being the selection of the case study.

**Table 1.**
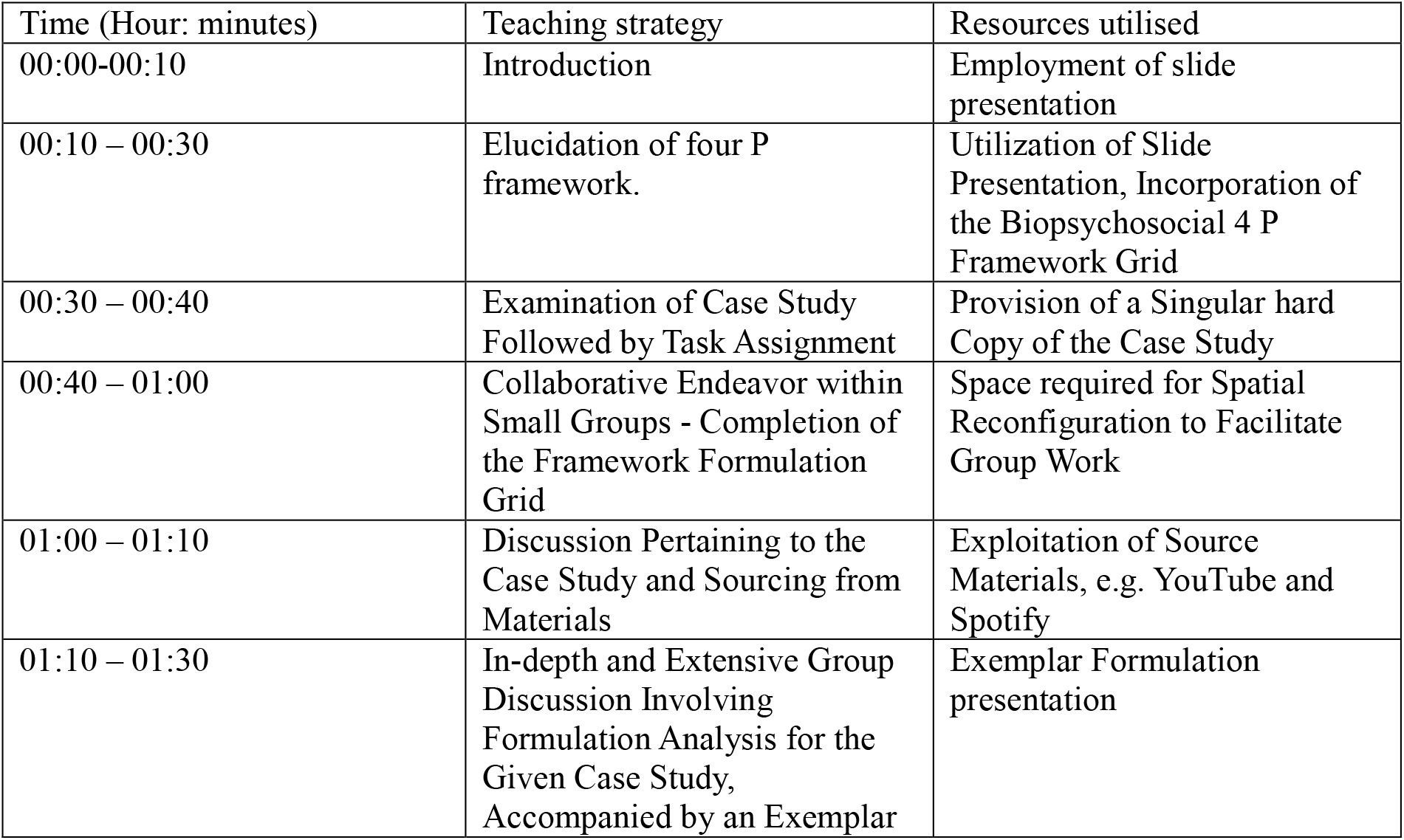
Structure of the psychiatric formulation tutorial.

### Recruitment

Following the tutorial, a survey was dispatched to all students who had attended the tutorial via a gatekeeper utilizing their respective student email addresses. The email comprised consent forms for both the survey and potential involvement in subsequent interviews. A link to the survey, facilitated through Qualtrics, was also incorporated. Participation by the students was voluntary, with no incentives provided. Anonymity was upheld throughout the survey process. Participation in both the survey and interviews was not mutually exclusive.

### Data collection

Virtual interviews were conducted using Microsoft Teams software, with sessions being recorded and transcribed. The transcriptions underwent review and correction by researchers DH and JMCF. The corrected transcriptions were subsequently returned to participating students for the purpose of triangulation. Instances that were not amenable to transcription or that exhibited ambiguity in content or meaning were singled out and relayed back to the participants for further clarification. Any points that remained unclear or incomprehensible were consequently excluded from the ensuing analysis.

### Data analysis

Data from the survey was transferred to microsoft word to develop charts to visually aid interpretationof of the data.

The transcribed data fomr the interviews underwent successive readings by the research team (DH, JMCF) to cultivate familiarity and subsequent data analysis process followed an adapted six-stage guide (Braun et al., 2006). Initial codes were formulated, and potential themes were deliberated upon until a consensus was reached. Microsoft Word software was employed for manual color coding of the data. Themes and subthemes were continuously reviewed and subject to discussions, thus aligning them optimally with the entirety of the dataset.

## Results

### Survey data

Two questions from the survey emerge as pertinent for the results section (Question 1 and Question 2). The remaining questions were not deemed relevant to the discussion and are thus excluded from the results.

Both Question 1 and Question 2 yielded highly positive outcomes. The X-axis identifies three distinct case studies (Kurt Cobain, Amy Winehouse, and Kanye West) (see Figure 1 and Figure 2). No statistically significant differences between the case studies were observed. In the Kurt Cobain tutorial, 88% of attending students responded positively to Question 1, indicating a favourable impact on their engagement compared to anonymous or fictional cases. Additionally, 93.7% answered positively to Question 2, expressing that future celebrity references would enhance their learning (n=17). Similarly, in the Amy Winehouse tutorial, 100% responded positively to Question 2, with 88% also positively reacting to Question 1 (n=16). In the case of the Kanye West tutorial, which had notably higher participation, 95% provided positive responses to both Question 1 and Question 2 (n=44).

**Figure 1.**
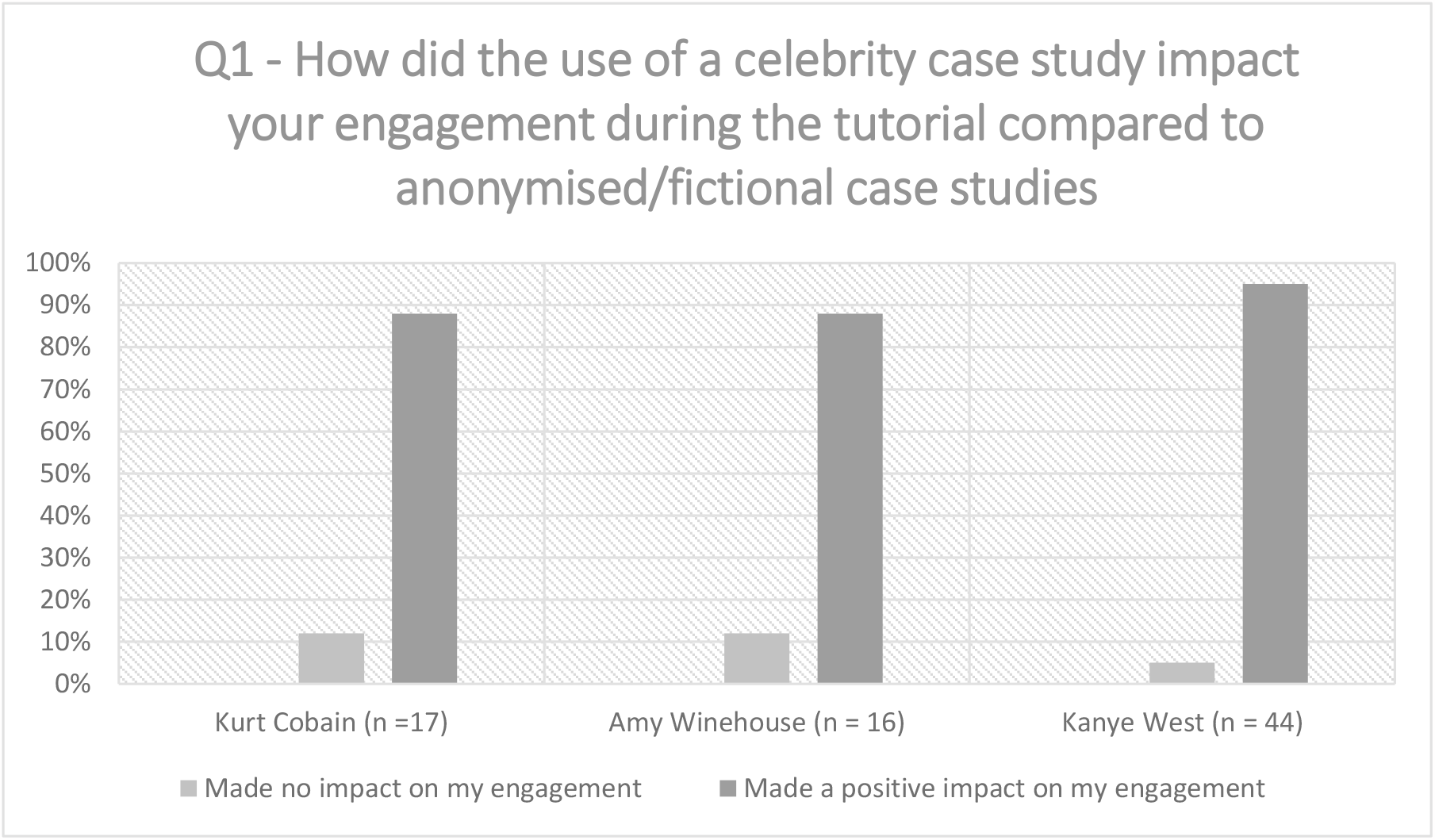
Quantitative data from survey outlines the impact of using the celebrity narrative compared to an anonymised case.

**Figure 2.**
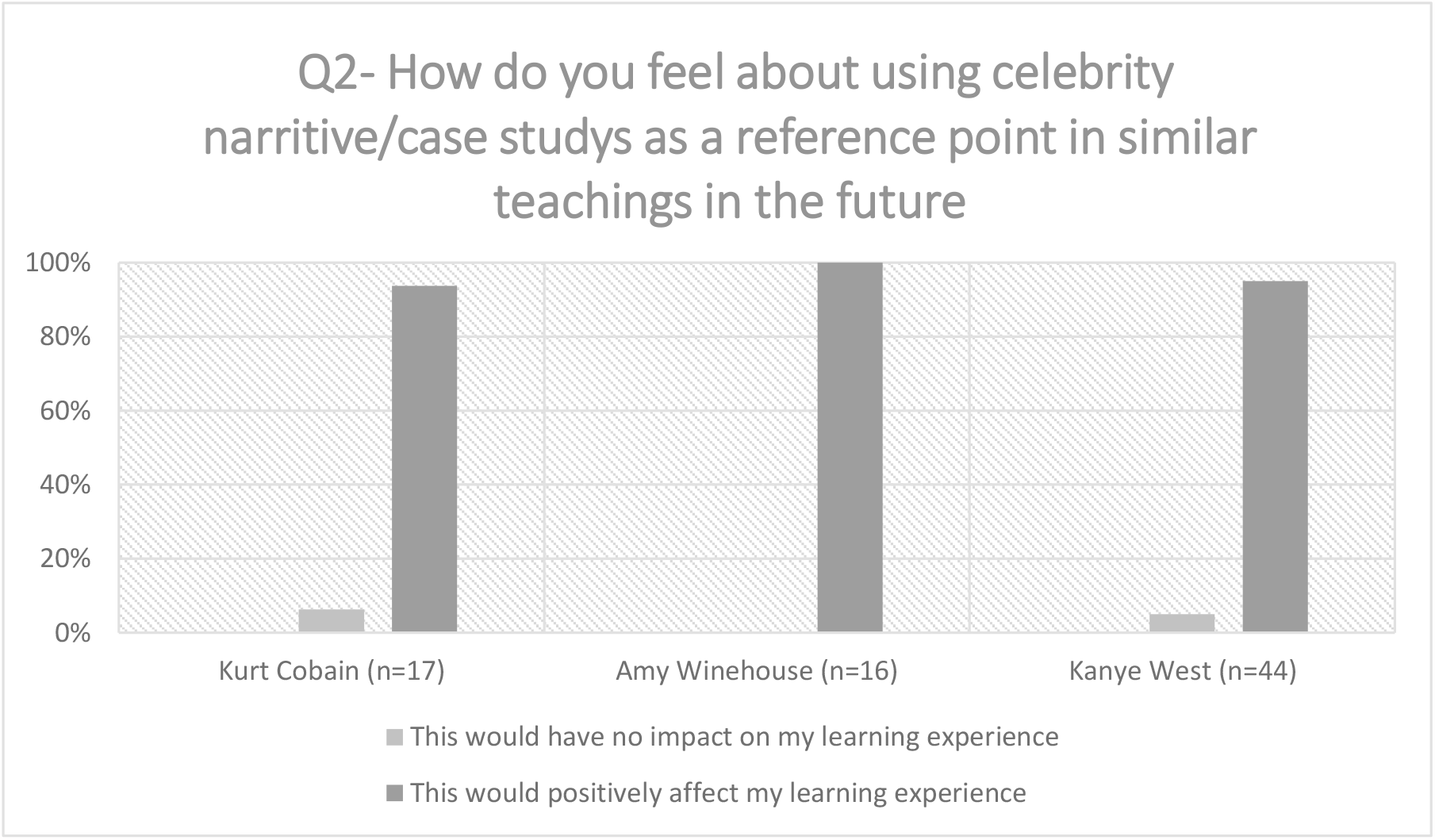
Quantitative data from survey, outlines the students’ feelings about using similar celebrity-based case studies in the future.

### Qualitative data

Four major themes and twelve corresponding subthemes emerged from the interviews (see Table 2).

**Table 2.** Themes, and corresponding subthemes generated from interviews.

| Themes | Subthemes |
| --- | --- |
| 1. Understanding of biopsychosocial formulation | 1. Foundational understanding and discernment of clinical applicability<br>2. Contextualisation of knowledge surrounding interplay of biological, psychological, and social factors.<br>3. Limitations in reductionist biomedical models |
| 2. Cognitive engagement | 4. Pedagogical constructivism<br>5. Universality and generalisability<br>6. Facilitation of reflective cognitive processes |
| 3. Emotional salience | 7. Secure and enjoyable learning environment<br>8. Humanisation of celebrity figures<br>9. Affective learning |
| 4. Ethical considerations | 10. Sensitivity and confidentiality<br>11. Objectivity and impartiality in dissemination of information<br>12. Perception of educational benefit |

### 1 Understanding of biopsychosocial formulation

#### 1.1 Foundational understanding and discernment of clinical applicability

A core understanding of formulation structure and prognostic implications was evident:

“*It was impactful in terms of getting across a biopsychosocial formulation… It puts things very nicely into boxes… You’re doing it for whatever else comes down the road*.. *You link the importance of doing the formulation an awful lot more” (Student 4)*.

The tutorial assisted students in recognising the importance of good formulation skills in clinical practice:

*“You recognise the formulation is an important piece of work to be able to do”* (Student 7).

Students expressed their experiences of reflecting on the application of the formulation during their attachments:

*“I’m trying to be a sponge and tyring to appreciate what’s going on… it’s not until you sit down and try to do the formulation or find a really good formulation in the notes*.*… you recognise that the formulation is an important piece of work…*.*” (*Student 2).

#### 1.2 Contextualisation of knowledge surrounding interplay of biological, psychological, and social factors

Several students acknowledged intrinsic complexity of the formulation.

‘*It’s a tricky thing to learn…. there are so many moving factors’’* (Student 11)

Students communicated an appreciation of how several multifactorial influences can impact a psychiatric presentation and described how their own perceptions have changed:

*“You have preconceptions… You could actually begin to understand it a bit more as to what would happen to the person and why it was happening” (*Student 1).

#### 1.3 Limitations in reductionist biomedical diagnostic models

Students displayed multiaxial cognitions, considering processes as opposed to cross sectional events: “*There was a whole process before the suicide to consider” (Student 4)*.

Students began to appreciate the importance of social information often not considered in reductionist diagnostic models:

*“Like there was so much going on with her that you wouldn’t have known…. everything else that went down… in her childhood, family life… all of that”* (Student 8).

### 2 Cognitive engagement

#### 2.4 Pedagogical constructivism

Students described clearly the benefits having a baseline knowledge of the celebrity and the general narrative:

“*It makes a power of a difference”* (Student 1).

“*Coming in with a baseline knowledge and building from that… It’s left me with more of a lasting impression than someone who’s fictitious’ (Student 3)*.

#### 2.5 Universality and generalisability

Students described the significance of universal knowledge describing it as *“really important”* (Student 14). They appreciated that more niche personal interests may not benefit the group on the whole and appreciated the balance between clinical content and choice of celebrity narrative:

*“Picking someone like a random footballer that would benefit me, that cuts off another segment of people in the class” (Student 3)*.

*‘I don’t think the choice of celebrity is vital, I would engage with the case on its own merit’*.

#### 2.6 Facilitation of reflective cognitive processes

Engagement continued post-tutorial in a collaborative non-formal capacity. Notably it also promoted reflective thoughts on art attributed to the celebrity:

“*It generated conversation with my friends”* (Student 7)

“*When I’m listening to his songs…. the lyrics… they have more meaning now’*” (Student 1).

### 3 Emotional salience

#### 3.7 Collaborative learning environment

The invoked emotions promoted a shift in the manner of discourse. It also promoted a healthy educational environment as students acknowledged the interest of their peers:

“*People were talking about it*.. *more empathetically*.. *compared to an anonymised cases” (Student 9)*.

“*It encouraged an awful lot of debate among people”* (Student 5).

*“I got a sense that everybody was more interested and invested”* (Student 17).

#### 3.8 Humanisation of celebrity figures

Many students felt as opposed to further sensationalising a celebrity narrative their enhanced understanding of the narrative fostered positive emotions:

*“If anything, it breaks down barriers’* (Student 14).

*“I think the tutorial…. It humanises them more… gives them more personhood’* (Student 1).

#### 3.9 Affective learning

Students discussed a range of emotions and the potential impact these emotions may have on their future practices,

*“A greater emotional response because they it’s not just connecting to someone who’s real, it’s*

*connecting to someone who’s not only real, but they have connected to before in a way”* (Student 2).

*“We connected with them before… something that’s really personal. And I think that’s also why it really pushes the emotional attachment’* (Student 13)

*“Guilt, I had an opinion of her prior without known all the details*.. *you can’t make any opinions of anyone without knowing the details… and we might take that into our careers, not to judge someone until you know all the facts”* (Student 7)

### 4 Ethical considerations

#### 4.10 Sensitivity and confidentiality of information

Students unanimously reported no concerns regarding the handling or distribution of sensitive information:

*“I found it the opposite of insensitive, it was informative”* (Student 6).

*“We were all respectful in our use of information”*. (Student 4)

#### 4.11 Objectivity and impartiality in the dissemination of information

Students appreciated a professional approach to the tutorial with objective, impartial information to reduce or eliminate ethical objections to sharing sensitive information,

“*It was done very respectfully*… *I didn’t find it insensitive”* (Student 5). “*Just dealing in facts… Things were quite black and white”* (Student 12)

#### 4.12 Perception of educational benefit

Students acknowledge the privilege of societal position in the medical community and acknowledged the communal educational objective in the room.

*“As medical students… we might have a different sense of intimate details”* (Student 13).

*Everyone in the room was using that information for an educational purpose”* (Student 9).

## Discussion

The survey data’s outcomes exhibit considerable promise, indicating improved engagement levels among students when a celebrity-based case study is implemented compared to conventional anonymized or fictitious case-based learning paradigms. Moreover, there is a prevailing sentiment among students that forthcoming instructional approaches employing this methodology will yield positive impacts on their learning experiences. The absence of discernible variations in responses across distinct case studies further supports the notion that these findings may possess transferability across other celebrity figures, possibly contingent upon the attainment of a certain threshold of public prominence.

Through qualitative analysis, twelve subthemes have emerged, distributed across four overarching themes. These themes have exhibited minimal divergence among the three case studies and five interviews. The principal themes encompass comprehension of formulation, cognitive engagement, emotional significance, and ethical deliberations.

### 1 Understanding of formulation

#### 1.1 Foundational understanding and discernment of clinical applicability

The findings of this study suggest the viability of employing a celebrity narrative to cultivate a foundational understanding of psychiatric formulation among final year medical students.

The attainment of a fundamental understanding is the most attainable objective among medical students given inherent time constraints in this cohort. Attaining advanced skills in this area is a pursuit more aligned with the expectations of advanced psychiatric trainees to whom this study is not directly concerned. The results suggest that the establishment of a rudimentary cognitive framework of formulation of organising knowledge and appreciating value is achievable using the methodology outlined in this study. The implication of attaining this core understanding is that it may underwrite the progression along the continuum of knowledge acquisition towards synthesis and evaluation as outlined by (Bloom et al., 1956) and (Anderson & Kratwohl, 2001) respectively. Devoid of this foundational comprehension, the progression along the cognitive continuum becomes an onerous task for the student.

Furthermore, the students appreciated that this newly acquired knowledge base will permit them to comprehend and engage with documented formulations under the supervision of an experienced consultant psychiatrist during clinical attachments. Attaining this pre-requisite knowledge allows for more profound situational and experiential learning experiences during clinical attachments (Lave & Wenger., 1991; Yardley et al., 2012). The self-identification of existing knowledge and corresponding gaps highlights advantageous metacognitive states as delineated by Flavell (1979). Students acknowledged their new understanding might act as a cognitive conduit linking their own knowledge to those of an experienced consultant psychiatrist. This linkage enhances the student’s capacity to apprehend the intricate processes underpinning the development of sophisticated biopsychosocial management strategies and is compatible with some of the pedagogical processes outlined in mastery learning (MacNamara & Maitra, 2019).

#### 1.2 Contextualisation of the interplay between biological, psychological, and social factors. 1.3 Limitations in reductionist biomedical models

Beyond the fundamental grasp of formulation, students exhibited engagement with more advanced cognitions. The intervention prompted students to analyse the intricate multifactorial nature of an individual’s mental health, encompassing the interaction between intricate social elements and how that shape an individual’s cognition and eventually impacts on presentation. The appreciation of moving factors and acknowledgement of changing perceptions of individuals indicates attainment of an understanding of the complex interplay of biopsychosocial influences.

Several students demonstrated a nuanced contextualization of dynamic social factors, the implications on formulation design and subsequent effects on presentation and prognosis. This appreciation for non-clinical information surpasses the confines of the standard biomedical reductionist diagnostic model.

### 2 Cognitive engagement

#### 2.4 Facilitating pedagogical constructivism and 2.5 Universality and generalisability

The study highlights the significance of constructivism, a pedagogical framework outlined by Vygotsky (1978). Through the employed methodological approach, the constructivist pedagogical process was nurtured, facilitating rapid student engagement, and emphasizing the universal applicability of the celebrity case study. This approach augmented participation rates and enriched collaborative discourse, potentially fostering a conducive milieu for social learning (Bandura, 1977). The study’s findings align with advantages observed in a flipped classroom approach that involves pre-tutorial work (Tolks et al., 2016). Notably, student compliance with pre-tutorial work in undergraduate education is estimated to be 30% (Bhavsar, 2020; Hattenberg & Steffy, 2013; Hoeft, 2012). It is conceivable that an existing communal knowledge of a notable celebrity would far surpass this figure, offering a shared knowledge base and potentially enhancing in-class engagement and participation reaping some of the advantageous of a flipped classroom.

The chosen celebrity case study’s universality and generalizability were recognized by students as crucial factors. Identifying renowned celebrity figures resonating with a diverse class cohort, while embodying essential clinical content, was acknowledged as intricate yet valuable. Balancing classroom dynamics to engage the entire class was emphasized, ensuring inclusivity. The study underscores the potential impact of case study choice, universality, and generalizability on the learning environment and promotion of social learning processes. While the synergy between unregulated pre-existing knowledge and structured pre-tutorial assignments requires further exploration, their concurrent use could yield positive outcomes.

#### 2.6 Facilitation of reflective processes

The study yielded notable results in cultivating and stimulating reflective practices across various dimensions. This heightened students’ awareness of personal and others’ attitudes, fostering metacognitive awareness. This led to self-directed exploration, including reevaluating media portrayals of the celebrity. Such reflective processes align with connectivism principles (Siemens, 2005), utilizing diverse sources and communication modes. While the study’s scope doesn’t fully explore the post-tutorial reflection’s advantages in enhancing formulation learning, routine art and media interaction after tutorials could potentially strengthen comprehension. This outcome is promising for engaging students who might otherwise lose interest in the subject matter.

### 3 Emotional salience

#### 3.7 Secure and enjoyable learning environment

No negative emotions towards formulation were found in this study, fostering a positive learning environment. Notably, ‘overwhelm’ and ‘daunt’ feelings hinder psychiatric trainees’ engagement (Bagster, 2021). This study’s outcomes suggest its methodology’s pedagogical mechanisms curbed adverse emotional responses. Plausible theoretical justifications elucidated from results include positive collaborative learning environment establishment, cognitive load mitigation through prior knowledge or impartial information dispensation.

#### 3.8 Humanisation of celebrity figures

Students perceived an increased sense of authenticity with celebrity case studies, fostering engagement akin to real clinical encounters. Contrary to concerns about sensationalism and limited generalizability, results suggest celebrities can be humanised, nurturing empathy and personal connection resembling genuine clinical interactions. This resonates with humanistic learning theory (DeCarvalho, 1991). Analysing the methodology’s intricate processes for heightened emotional salience poses challenges. Nevertheless, the study’s approach holds potential to evoke heightened emotional experiences. Students exhibited empathy and emotional connection, with one reflecting on their feelings of guilt and its impact on future practice. This outcome resonates with the profound impact emotion can have on learning described by Bloom, (1956).

### 4 Ethical considerations

#### 4:9 Sensitivity and confidentiality, 4.10 Impartiality and objectivity of information

Ethical concerns arise from using sensitive public information about well-known figures without explicit consent. The ‘Goldwater rule’ in APA’s ethical guidelines aims to protect both psychiatry’s integrity and public figures from psychoanalytic scrutiny (APA, 2009). Critiques propose that this rule is too restrictive for modern practice (Kroll & Pouncey, 2006), that it should not hinder scholarly pursuits (Appelbaum, 2017) and that the fostering of sensitive discussions may be advantageous (Friedman, 2008). Despite apprehensions about exacerbating celebrity culture, Walsh (2005) ceased using celebrity narratives in pedagogy due to information sensitivity and perpetuation of unwarranted celebrity status. Results in this study indicate these concerns should not deter future engagement. The methodology nurtured empathetic feelings, delivering impartial information for comprehension and humanizing celebrities, counteracting media sensationalism. This emphasizes the viability of this approach in education.

#### 4.11 Perception of educational benefit

The outcomes gleaned from the students’ perspectives on these ethical considerations present a novel dimension, which add to the ethical discourse. Two significant discussion points emerge from the results here. First students are aware of their entitlement to sensitive and confidential information given their position as future health care professional and the studies methodology may have reinforced that awareness. Secondly, students appreciated the communal educational goal within the confines of the tutorial, that the purpose was education not speculation. The results bear significance as they suggest students are conscious to the possibility of the existence of an *‘educational license’*, which may bend around ethical concerns akin to the way an artistic license may deviate around conformity.

## Conclusion

This study presents an initial exploration of utilising celebrity narratives as a pedagogical tool for teaching psychiatric formulation to medical students. This approach proves feasible in enhancing fundamental understanding and contextual insights while mitigating negative attitudes and perceptions. It also raises ethical considerations and fosters cognitive and emotional engagement. The recurring themes across diverse cases highlight the approach’s robustness. Findings could guide cost-effective curriculum development for early biopsychosocial formulation education. Future research might investigate the prolonged impact on attitudes and skills on the students who proceed to become psychiatric trainees. Lastly, this study’s preliminary exploration into pedagogical processes could serve as a foundation for future research into harnessing human fascination with celebrities for medical education.

### Limitations

- The interview participant count was limited, although indications of data saturation were observed.
- Not all tutorial attendees aspire to pursue careers in psychiatry. Volunteers for interviews might over-represent those interested in psychiatry. This specific subset was not collected.
- Quantifiable improvement in formulation knowledge or skill was not assessed, relying on discussion analysis to infer understanding. This may introduce subjective interpretation due to the informal interview format.

## Data Availability

All data produced in the present study are available upon reasonable request to the authors

